# Development and Validation of Natural Language Processing Algorithms in the ENACT National Electronic Health Record Research Network

**DOI:** 10.1101/2025.01.24.25321096

**Authors:** Yanshan Wang, Jordan Hilsman, Chenyu Li, Michele Morris, Paul M. Heider, Sunyang Fu, Min Ji Kwak, Andrew Wen, Joseph R Applegate, Liwei Wang, Elmer Bernstam, Hongfang Liu, Jack Chang, Daniel R. Harris, Alexandria Corbeau, Darren Henderson, John D Osborne, Richard E Kennedy, Nelly-Estefanie Garduno-Rapp, Justin F. Rousseau, Chao Yan, You Chen, Mayur B. Patel, Tyler J. Murphy, Bradley A. Malin, Chan Mi Park, Jungwei W. Fan, Sunghwan Sohn, Sandeep Pagali, Yifan Peng, Aman Pathak, Yonghui Wu, Zongqi Xia, Salvatore Loguercio, Steven E. Reis, Shyam Visweswaran

**Affiliations:** Clinical and Translational Science Institute, University of Pittsburgh, Pittsburgh, PA, USA; Department of Health Information Management, University of Pittsburgh, Pittsburgh, PA, USA; Department of Biomedical Informatics, University of Pittsburgh, Pittsburgh, PA, USA; Biomedical Informatics Center and Department of Public Health Sciences, Medical University of South Carolina, Charleston, SC, USA; McWilliams School of Biomedical Informatics, University of Texas Health Science Center at Houston, Houston, TX, USA; McGovern Medical School, University of Texas Health Science Center at Houston, Houston, TX, USA; Division of General Internal Medicine, McGovern Medical School, University of Texas Health Science Center at Houston, Houston, TX, USA; Clinical and Translational Science Institute, University of Rochester Medical Center, Rochester, NY, USA; Institute for Biomedical Informatics, University of Kentucky, Lexington, KY, USA; Department of Biomedical Informatics and Data Science, University of Alabama at Birmingham, Birmingham, AL, USA; Division of Gerontology, Geriatrics, and Palliative Care, Department of Medicine, University of Alabama at Birmingham, Birmingham, AL, USA; Clinical Informatics Center, University of Texas Southwestern Medical Center, Dallas, TX, USA; Department of Neurology, University of Texas Southwestern Medical Center, Dallas, TX, USA; Department of Biomedical Informatics, Vanderbilt University Medical Center, Nashville, TN, USA; Department of Surgery, Vanderbilt University Medical Center, Nashville, TN, USA; Department of Gerontology, Hebrew SeniorLife, Marcus Institute for Aging Research, Boston, MA, USA; Department of Artificial Intelligence and Informatics, Mayo Clinic, Rochester, MN, USA; Center for Clinical and Translational Science, Mayo Clinic, Rochester, MN, USA; Department of Medicine, Mayo Clinic, Rochester, MN, USA; Department of Population Health Sciences, Weill Cornell Medicine, New York, NY, USA; Clinical & Translational Science Center, Weill Cornell Medicine, New York, NY, USA; Department of Health Outcomes and Biomedical Informatics, University of Florida, Gainesville, FL, USA; Department of Neurology, University of Pittsburgh, Pittsburgh, PA, USA; Scripps Research Translational Institute, Scripps Research, La Jolla, CA, USA

**Keywords:** translational research, electronic health records, natural language processing, network, enact

## Abstract

Electronic health record (EHR) data are a rich and invaluable source of real-world clinical information, enabling detailed insights into patient populations, treatment outcomes, and healthcare practices. The availability of large volumes of EHR data are critical for advancing translational research and developing innovative technologies such as artificial intelligence. The Evolve to Next-Gen Accrual to Clinical Trials (ENACT) network, established in 2015 with funding from the National Center for Advancing Translational Sciences (NCATS), aims to accelerate translational research by democratizing access to EHR data for all Clinical and Translational Science Awards (CTSA) hub investigators. The present ENACT network provides access to structured EHR data, enabling cohort discovery and translational research across the network. However, a substantial amount of critical information is contained in clinical narratives, and natural language processing (NLP) is required for extracting this information to support research. To address this need, the ENACT NLP Working Group was formed to make NLP-derived clinical information accessible and queryable across the network. This article describes the implementation and deployment of NLP infrastructure across ENACT. First, we describe the formation and goals of the Working Group, the practices and logistics involved in implementation and deployment, and the specific NLP tools and technologies utilized. Then, we describe how we extended the ENACT ontology to standardize and query NLP-derived data, as well as how we conducted multisite evaluations of the NLP algorithms. Finally, we reflect on the experience and lessons learnt, which may be useful for other national data networks that are deploying NLP to unlock the potential of clinical text for research.

## Introduction

Electronic health record (EHR) data serve as a rich and invaluable source of real-world clinical information, enabling researchers and healthcare professionals to gain comprehensive insights into patient populations, treatment outcomes, and healthcare practices^1^. By capturing a broad spectrum of clinical information, including demographic details, diagnosis, procedures, medications, laboratory test results, and clinical notes, EHR systems create a longitudinal record that mirrors the complexity and heterogeneity of modern healthcare. The accessibility of EHR data is paramount for advancing translational research and the application of cutting-edge technologies, including artificial intelligence and machine learning. Furthermore, these computational tools depend on robust, standardized, and interoperable EHR datasets to enable predictive modeling^2^, automated phenotyping^3^, risk stratification^4^, and decision support systems^5^ that can enhance clinical effectiveness, improve patient safety^5,6^, and ultimately shape the future of healthcare delivery.

The national Evolve to Next-Gen Accrual to Clinical Trials (ENACT) network^1^ was established in 2015 as the Accrual to Clinical Trials (ACT) network to enable cohort discovery from EHR data. This federated network connects EHR data repositories across 57 Clinical and Translational Science Awards (CTSA) hubs, enabling investigators to query the data of more than 142 million patients across the hubs (sites). The ENACT network integrates local Informatics for Integrating Biology at the Bedside (i2b2)^7^ and Observational Medical Outcomes Partnership (OMOP)^8^ data repositories (and eventually PCORnet^9^ data repositories) through the Shared Health Research Information Network (SHRINE) platform, which enables interactive querying of the data^10^. The network’s data, encompassing structured EHR information on demographics, diagnoses, procedures, medications, laboratory test results, and visits, extends back at least a decade, with some sites providing data for up to two decades. Updates to the data occur at least monthly.

The ACT network’s purpose was to enable national cohort discovery, particularly for multisite research such as clinical trials, including those supported by the Trial Innovation Network (TIN)^11^. However, the ENACT network’s goal is broader, including large-scale clinical and translational research using patient counts, distributed analytics, and ephemeral analytics enclaves. Furthermore, ENACT provides prep-to-research data, enables the generation of evidence for clinical decision-making, and serves as a resource for educating trainees.

The current ENACT network provides access to structured EHR data, enabling significant advances in cohort discovery and research across this national network. However, while structured EHR data offers valuable insights, much clinical information remains embedded within unstructured EHR data. The unstructured EHR data, including clinical encounter notes, radiology reports, pathology reports, and other narrative documents, are challenging to analyze due to their free-text format. To harness the full potential of EHRs for translational research, applying natural language processing (NLP) to extract research-usable data from clinical notes is essential. Recognizing this critical need, the ENACT NLP Working Group was established to make NLP-derived data accessible and queryable across the network. Such data will enhance the analytical capacity of the network, enabling researchers to tap into previously inaccessible information in clinical notes and generate new insights that can drive advances in translational research and clinical care.

In this article, we provide a comprehensive overview of the development and deployment of NLP infrastructure in ENACT. We describe the formation and goals of the Working Group, the policies and logistics involved, and the specific NLP algorithms and tools utilized. We also describe the extension of the ENACT ontology to standardize and query NLP-derived data across the network. Furthermore, we provide a practical guide on multisite evaluation of NLP algorithms, highlighting their performance, scalability, and adaptability across diverse healthcare systems. We also include an in-depth reflection on the experiences and lessons learned from this journey, which may be useful for other national data networks, such as the PCORnet^9^ and the All of Us Research Program^12^, which are interested in using NLP to unlock the potential of clinical notes for research.

## Results

### The ENACT NLP Working Group

We launched the ENACT NLP Working Group with participation from thirteen ENACT sites in 2023 (see Figure 1). Each of the participating sites had a ready source of clinical notes (e.g., a clinical data warehouse (CDW) or an Epic Clarity reporting database), existing information technology (IT) infrastructure for computation, and some NLP expertise. Furthermore, with support from the local CTSA hub leadership, the sites could obtain clinical notes, access computing to implement NLP algorithms and manage other participation logistics. This enabled the Working Group to develop guidelines and implement NLP algorithms collaboratively and rapidly. Regular virtual meetings and a dedicated Slack workspace facilitated discussions and communication among members of the Working Group.

**Figure 1.**
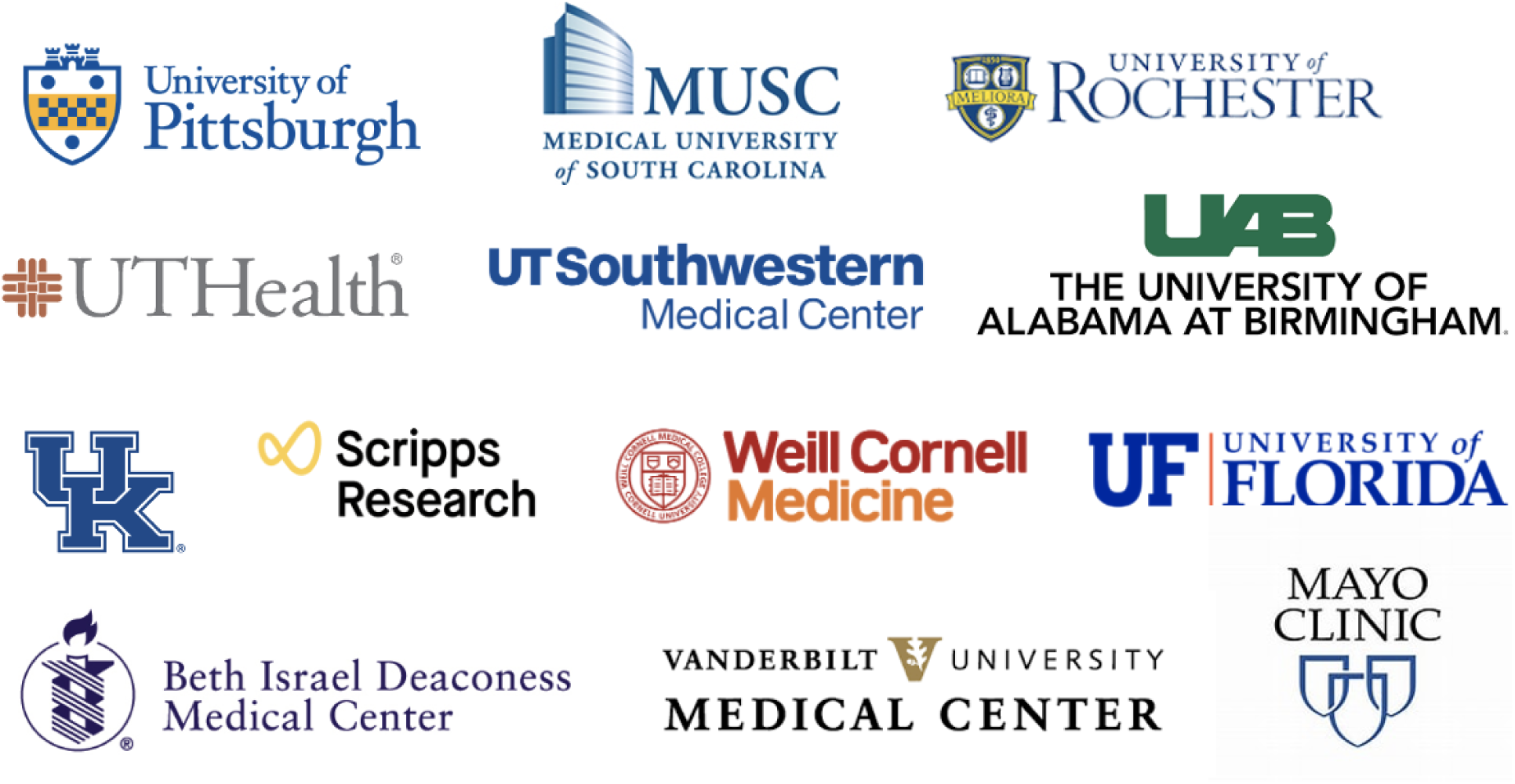
Participating sites in the ENACT NLP Working Group.

During the initial discussions, we discovered two challenges: 1) Processing all clinical notes at each site (which could number in the millions) and extracting every biomedical entity from each note (which could number in the tens of thousands) was infeasible, and 2) Deploying a general-purpose NLP algorithm capable of extracting all entities proved unrealistic and unlikely to perform optimally. Instead, specialized NLP algorithms targeting specific entities for particular tasks with greater precision were deemed more practical. Further, to maximize the limited funding available to the Working Group, we divided the participating sites into five focus groups, each targeting a specific task in the context of a disease or condition. Each focus group consisted of two development sites and at least two additional validation sites. The development sites were tasked with jointly designing and validating a specialized NLP algorithm, while the validation sites were responsible for evaluating the algorithm. After validation, the NLP algorithm can be deployed across the entire network. Some development sites leveraged already-funded local projects or focused on an algorithm already in development for an ongoing project. This strategy significantly reduced the resources and effort needed to develop and deploy several algorithms. **Table 1** lists the focus groups and associated development sites and validation sites.

**Table 1.**
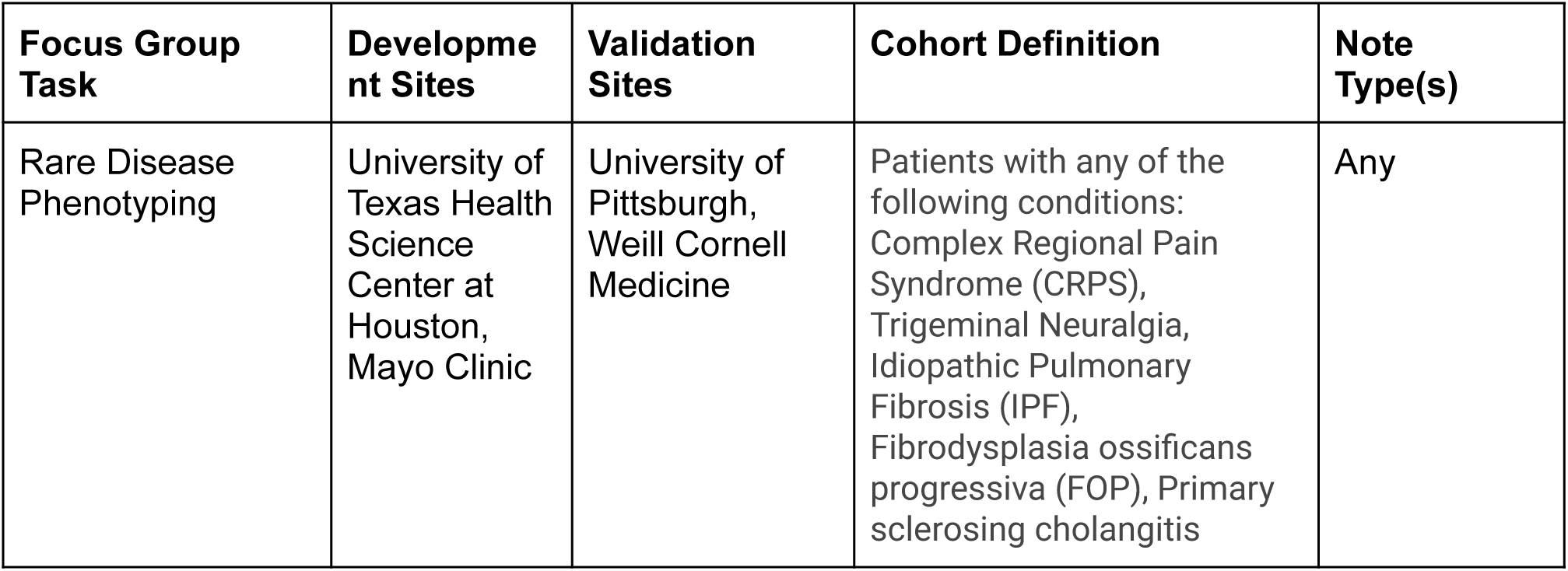
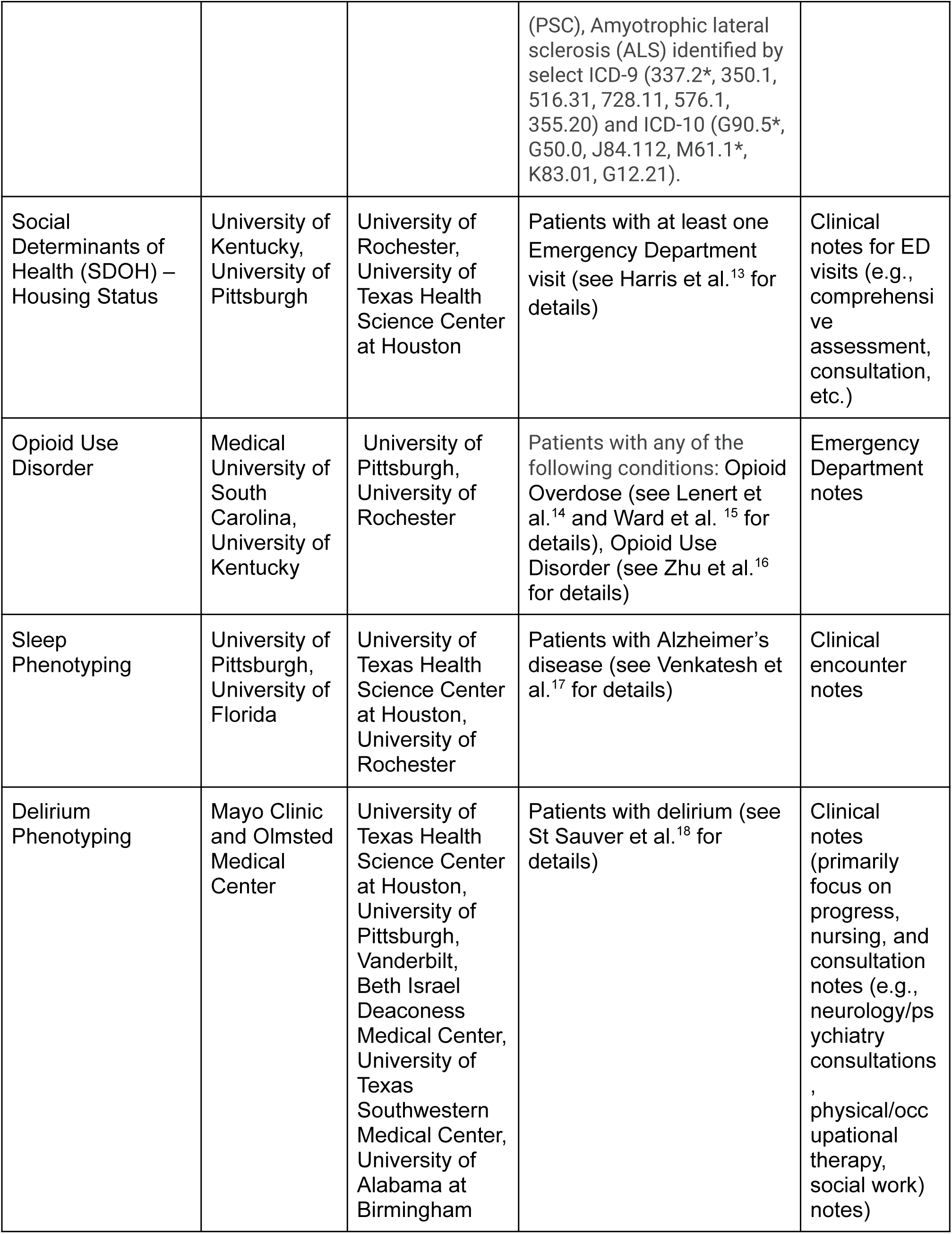
Focus group tasks and associated development sites, deployment sites, cohort definitions, and clinical note types.

During later discussions, we realized that designing a specialized NLP algorithm targeted for a task required specifying a patient cohort and a particular type of clinical note. For example, for the SDOH – Housing Status task, the patient cohort included individuals with substance use disorder (specifically those with stimulant and opioid use disorders with ICD-10-CM diagnosis codes of F11.*, F14.*, F15.*, T40.*, and T43.6*), and EHR data was limited to emergency department notes. The algorithm would not be expected to be applied to other types of patients or notes. Thus, we required each focus group to provide a clear cohort definition and identify the type of note for the development of the NLP algorithm. **Table 1** provides the cohort definitions and clinical note types identified by each focus group.

### Technologies

During this process, the NLP working group has worked collaboratively on selecting and implementing NLP tools for extracting medical concepts from clinical notes. The team also extended the ENACT common data model (CDM) to incorporate NLP-derived data while maintaining flexibility for different projects. Standardized conventions for storing NLP-extracted entities and contextual attributes were established, ensuring seamless integration with structured EHR data. Additionally, new ontologies were developed to facilitate querying

NLP-derived concepts across the national EHR network, and a federated evaluation framework was introduced for cross-site validation of NLP algorithms. To enhance reliability, the team also implemented a standardized error analysis process, utilizing an established taxonomy to refine model performance and assess generalizability across institutions. These collective efforts streamlined clinical textual analytical capabilities within the ENACT network. Details about these technology details can be found in the Method section.

## Overview of the ENACT NLP Workflow

**Figure 2** presents an overview of the ENACT NLP workflow that has been developed by the Working Group, which is described step by step below.

**Figure 2.**
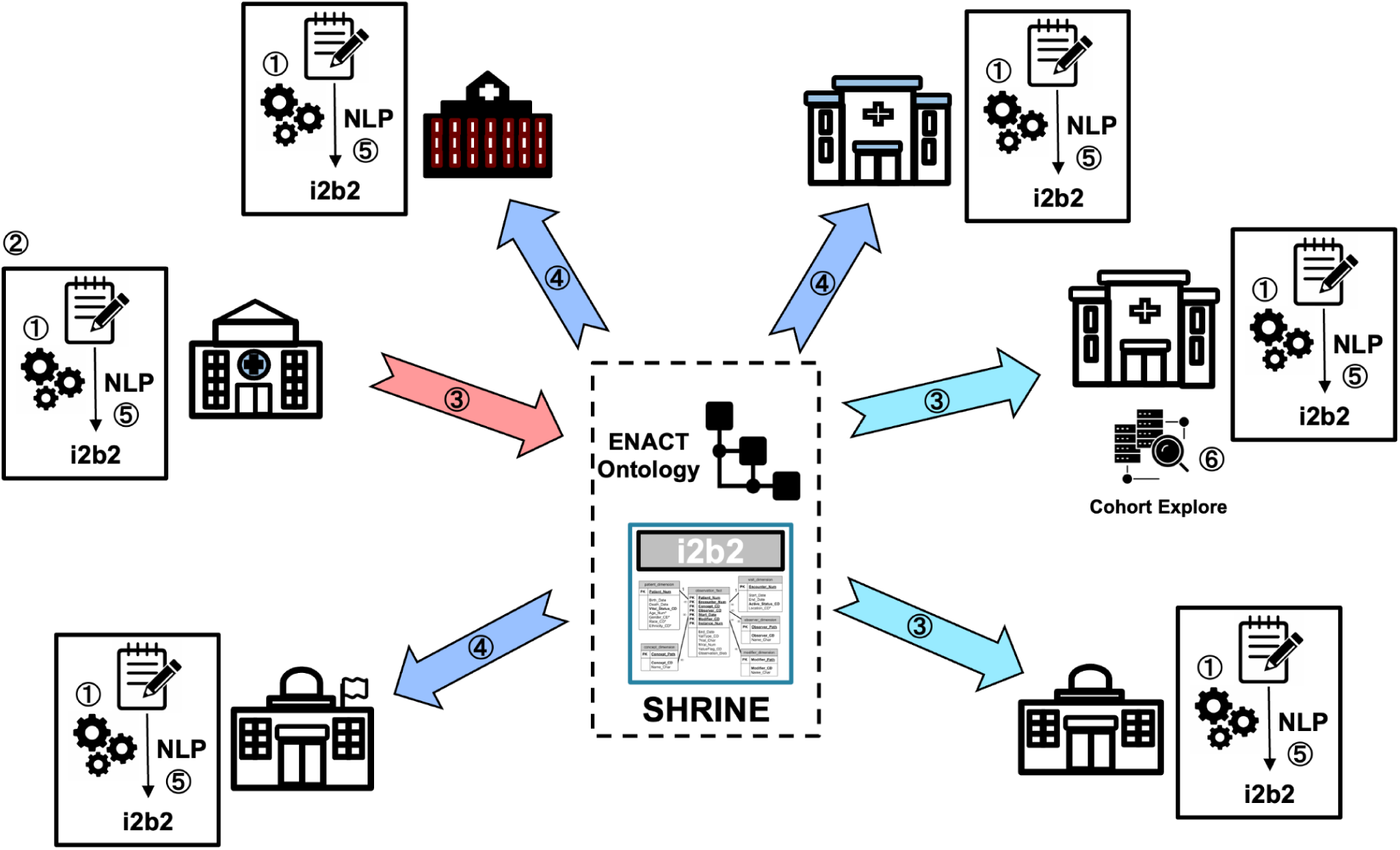
An overview of the ENACT NLP workflow.

① NLP Infrastructure: Each site participating in the NLP initiative will establish a source of clinical notes (such as a CDW or Epic’s Clarity reporting database), allocate computing resources for NLP processing, and deploy the OHNLP Toolkit. In addition to the ENACT Institutional Review Board (IRB) approval for structured EHR data, obtaining clinical notes may require additional IRB approval.

② Algorithm Development: A site proposes and develops a new NLP algorithm, then contacts the Working Group to coordinate its validation and to partner with the Data Harmonization Working Group to create the ENACT ontology extension necessary for deploying the algorithm.

③ Algorithm Validation: The site that developed the algorithm will share it with validation sites, along with the associated specifications, such as the cohort definition, note type, and process for establishing the gold and silver reference standards, as directed by the Working Group.

④ Dissemination: Following validation, the Working Group will disseminate the algorithm and associated specifications to the ENACT network via an online repository such as GitHub. In addition, the Working Group will coordinate with the Data Harmonization Working Group and the Network Operations Working Group to deploy the ENACT ontology extensions across the network.

⑤ Site Integration: Each participating site will download the algorithm and associated specifications from the online repository, integrate it into their local NLP infrastructure, and populate the local ENACT data repository’s i2b2 observation_fact table with NLP-derived data. Furthermore, the site will update the ENACT ontology to include the extension required for querying NLP-derived data.

⑥ Investigator Use: Any ENACT investigator at any participating site will be able to use the SHRINE interface to create queries that can search NLP-derived data as well as structured

EHR data. The query will return patient numbers from participating sites in the same way as existing structured EHR data queries do.

This workflow, which includes infrastructure needs, algorithmic creation and validation, network-wide distribution, and local integration, provides a structured approach for introducing and scaling NLP capability in the network.

### Demonstration Projects

In this section, we present four demonstration projects, each representing the work of a focus group and showcasing a distinct area of research. Each project is at a different stage of development, reflecting variations in goals, challenges, and resource availability. While some focus groups have made significant progress and are close to integrating NLP-derived data into the network, others are in the early stages, concentrating on foundational tasks such as acquiring clinical notes, establishing NLP infrastructure, or refining the NLP algorithm. This variation underscores the dynamic and adaptive nature of the collaborative effort to develop and disseminate NLP capabilities across an extensive national network.

### Demonstration Project 1: Sleep Phenotyping Focus Group

The Sleep Phenotyping Focus Group is investigating sleep phenotyping within a cohort of Alzheimer’s Disease (AD) patients, using encounter notes in these patients. To extract relevant sleep phenotype information, we previously developed an NLP algorithm to extract key phenotypes such as snoring, napping, sleep problems, poor sleep quality, daytime sleepiness, nocturnal awakenings, sleep duration, and other nocturnal symptoms^29^. This project offers a structured approach for analyzing the sleep disturbances commonly observed in AD patients, ultimately contributing to a deeper understanding of their clinical implications.

The evaluation framework described earlier was applied to assess the NLP algorithm, and the performance is shown in **Table 2**. Two sites, University of Pittsburgh (Pitt) and the University of Florida (UF) are the development sites, while the University of Texas Health Science Center at Houston (UTH) is one of the validation sites (the additional validation site, the University of Rochester, is in the process of generating validation results). The algorithm’s behavior varied among the three sites. At Pitt, it had a high recall and low precision, indicating that it functioned with high sensitivity, whereas at UF and UTH, it had a low recall and high precision, indicating that it functioned more cautiously. Part of the reason for this discrepancy is that the data at Pitt is more evenly distributed across the sleep phenotypes, with sample sizes of 12 and 48 for 6 of the 9 concepts, whereas the data at UTH and UF is imbalanced in terms of sleep problems and sleep quality bad concepts, for which the algorithm generates many false negatives.

**Table 2.**
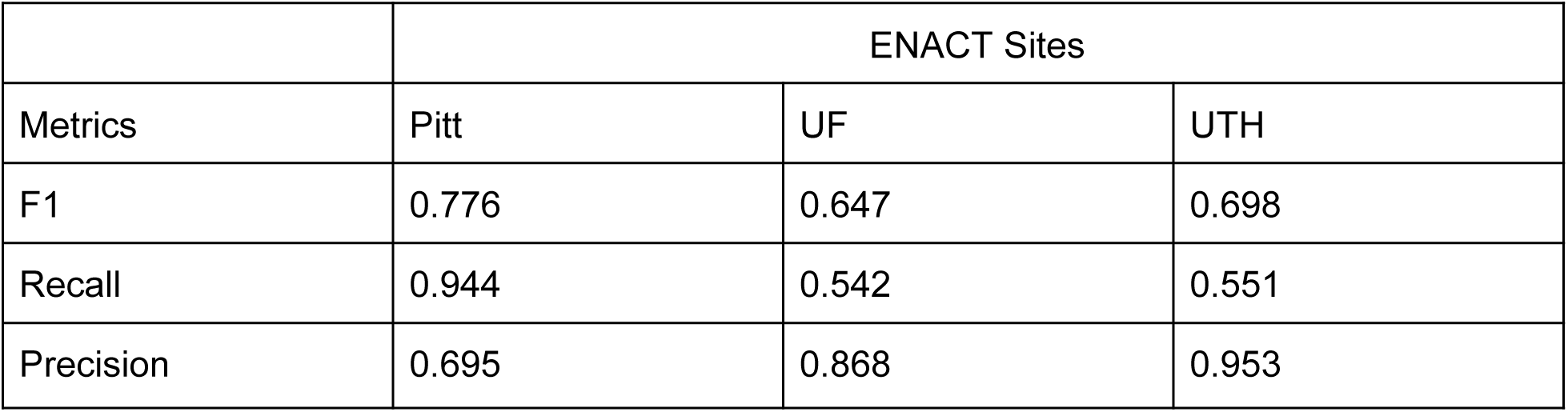
Performance of the algorithm developed by the Sleep Phenotyping Focus Group.

### Demonstration Project 2: Social Determinants of Health (SDOH) – Housing Status Focus Group

Housing is a key environmental social determinant of health (SDOH), closely associated with mortality and clinical outcomes. Housing Status Focus Group seeks to develop an NLP algorithm to extract the housing status of individuals from emergency department notes. This project aims to provide valuable insights into the impact of housing instability on health outcomes, thereby informing future interventions and support strategies.

The housing status NLP algorithm was developed to extract housing-related concepts such as homelessness, unstable housing, recovery housing, emergency housing, temporary housing, and exposure, and its performance is shown in **Table 3**. The University of Kentucky (UK) and Pitt are the development sites and University of Rochester (UR) and UTH are the validation sites. Each site created a gold standard for evaluation using a specific subset of patients treated in emergency departments; details of the NLP algorithm development and evaluation can be found in the relevant publication^13^.

**Table 3:**
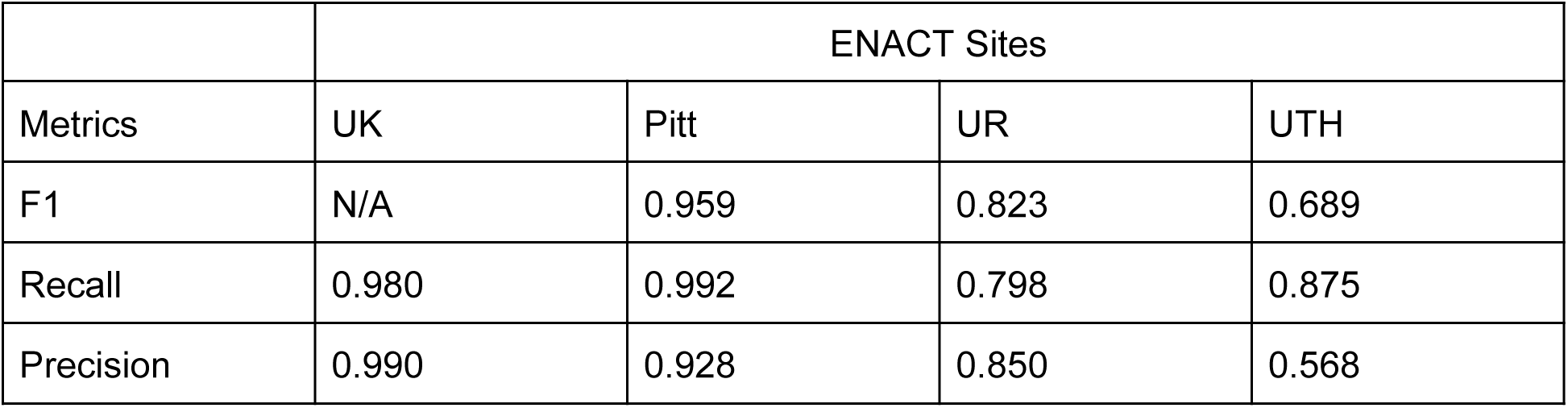
Performance of the algorithm developed by the Housing Status Focus Group.

### Demonstration Project 3: Opioid Use Disorder Focus Group

Patients who present to the Emergency Department with an opioid overdose (OOD) are at significant risk of death^30^. Identifying individuals with opioid use disorder (OUD) and at risk of OOD can aid in better treatment and counseling, particularly in the context of treating acute and chronic pain with opioids. ICD codes can be used for identifying OUD patients and those at risk of OOD, but they may not be available in the EHR at the time of the visit (Ward et al.^14^), and they are frequently absent in patients when evidence in their unstructured clinical notes indicates a risk for OUD(Zhu et al.^16^). In particular, Zhu et al. discovered that a lexicon-based strategy to identify patients at risk for OUD outperformed an ICD-based method to phenotype patients with OUD. The Opioid Use Disorder Focus Group is formalizing a phenotype based on the ICD code approach (Ward et al., Lenert et al., and Zhu et al.) so that it can be used as an initial silver standard for evaluation. The initial NLP phenotyping method will be based on Zhu et al.’s lexicon-based approach to identifying OUD, which will be refactored to work in the OHNLP framework. The NLP algorithm is currently being developed and validated across multiple sites.

### Demonstration Project 4: Delirium Phenotyping Focus Group

Delirium is a common geriatric syndrome characterized by an acute change in mental status, fluctuating course, lack of attention, and disorganized thinking or altered level of consciousness^31^. Accurate prediction of delirium could significantly improve patient outcomes through targeted interventions for hospitalized patients. For delirium case ascertainment, we will use Confusion Assessment Methods^32^, which is recommended by the NIDUS (Network for Investigation of Delirium: Unifying Scientists) network, as the gold standard for diagnosing delirium. NLP-CAM is an NLP-powered computational phenotyping tool that can identify a patient’s delirium status from EHR^33^. The tool was initially developed at the Mayo Clinic based on the evidence-based “confusion assessment method” (CAM), which includes 13 unique concepts (e.g., agitation, disorganized thinking, and fluctuation). These concepts range from neuropsychological characteristics to cognitive/memory complaints. We applied NLP-CAM to three test sites (UTHealth, UAB, and VUMC) and reported the out-of-box performance shown in **Table 4**. We observed moderate-high performance degradation due to institutional variations in the CAM screening, documentation, and patient characteristics. Our next step is to conduct federated refinement^34^ to optimize the NLP performance at each site.

**Table 4.** Performance of the algorithm developed by the Delirium Phenotyping Focus Group.

|  | ENACT Sites |  |  |  |
| --- | --- | --- | --- | --- |
| Metrics | Mayo | UTH | UAB | VUMC |
| F1 | 0.958 | 0.895 | 0.530 | 0.606 |
| Recall | 0.919 | 0.985 | 0.770 | 0.796 |
| Precision | 1.000 | 0.819 | 0.400 | 0.490 |

## Discussion

The ENACT NLP Working Group created NLP capability that is specifically suited to a multisite, federated network for supporting large-scale analytics. This capability enables the extraction of data from clinical narratives, facilitating research that involves collaboration across multiple sites, while simultaneously addressing data heterogeneity and scalability. Below, we highlight six areas where the NLP capability offers transformative potential in ENACT.

### Potential Applications

#### Clinical Trial Eligibility

The NLP capability will enhance the identification of participants for multisite clinical research, including clinical trials, by utilizing NLP-derived data with the existing structured data for eligibility criteria. By standardizing access to clinical narrative data through the extended ontology, ENACT will accelerate research recruitment across diverse populations, providing improved demographic representation that will enhance the generalizability of study findings. The capacity to identify eligible participants at this scale will significantly benefit multisite clinical research.

#### Federated Learning

The ENACT NLP infrastructure supports federated learning by enabling multisite collaboration in AI model development without centralizing sensitive patient data. Each participating institution can process unstructured EHR data locally to extract features or embeddings, which are then aggregated into global models. This approach allows ENACT NLP to leverage its broad network to create AI models that are generalizable across diverse patient populations and healthcare systems. By providing a scalable solution for distributed model training, ENACT NLP addresses the challenges of data heterogeneity and privacy in large-scale AI research.

#### Digital Twin Development

Digital twins, virtual representations of individual patients, are an emerging tool in precision medicine, simulating disease progression and treatment responses. Digital twin technology benefits significantly from the multisite capabilities of ENACT NLP. By extracting detailed and nuanced patient data from unstructured narratives across the network, ENACT NLP enables the creation of comprehensive digital twins that incorporate diverse clinical contexts. This large-scale infrastructure ensures that digital twins are representative of the heterogeneity observed across different healthcare systems and patient populations. These enhanced models support personalized simulations of disease progression and treatment outcomes, offering valuable insights for precision medicine on a national scale.

#### Public Health

ENACT NLP facilitates significant advancements in public health research by extracting population-level insights from clinical narratives. By analyzing unstructured data for disease patterns, healthcare utilization, and social determinants of health, ENACT NLP supports epidemiological studies and real-time surveillance of emerging public health threats. For instance, the system can identify trends in disease prevalence, monitor the spread of infectious diseases, and evaluate public health interventions across a diverse and extensive population. These capabilities enable targeted interventions, informed policymaking, and efficient resource allocation, addressing both acute and chronic public health challenges. This capability ensures that insights derived from unstructured data are both scalable and representative of the national population.

#### Multisite Large Cohort Studies

The integration of unstructured data into large cohort studies significantly enhances the granularity and scope of research. ENACT NLP enables the identification of complex phenotypes, such as those described in clinical narratives, which are often omitted in structured data alone. This capability is particularly valuable for studying rare diseases, as it allows researchers to pool data from multiple institutions to identify sufficient cases for robust analysis. By supporting large-scale phenotyping and longitudinal analyses, ENACT NLP facilitates cohort studies that can uncover complex relationships between clinical variables and outcomes, providing insights that would be unattainable with data from a single site.

#### Clinical Decision Support

The integration of NLP-derived insights into clinical workflows enhances decision-making processes by providing clinicians with comprehensive and actionable information. The multisite capabilities of ENACT NLP enhance clinical decision support systems (CDSS) by providing access to a vast and diverse range of clinical narratives from multiple institutions. ENACT NLP not only extracts actionable information from local unstructured EHR data, but also enables clinicians to identify similar patients across different healthcare systems within the network. For instance, clinicians can query ENACT NLP to find patients with comparable clinical profiles, including similar diagnoses, treatment histories, and demographic factors, from other institutions. This functionality allows clinicians to review past treatments and clinical outcomes for similar patients, offering valuable evidence to inform decision-making. By incorporating insights derived from a broader, multisite patient population, CDSS tools become more robust, relevant, and effective. This capability is particularly impactful in cases of rare or complex conditions, where local data may be insufficient to guide optimal care strategies.

ENACT NLP can also support other research areas by leveraging its multisite infrastructure to analyze unstructured clinical data. In precision medicine, ENACT NLP enables the extraction of patient-specific factors such as genetic variations and environmental exposures, facilitating personalized care through tailored treatments and predictive models. In quality improvement and healthcare operations, it helps identify workflow inefficiencies, care quality gaps, and patient safety issues by analyzing patterns in clinical narratives, driving improvements in operational processes and adherence to benchmarks. Additionally, ENACT NLP can advance health equity research by extracting data on social determinants of health (SDOH) from clinical notes, such as housing instability or food insecurity, enabling studies on healthcare disparities and informing equitable policy interventions. ENACT NLP can also play a critical role in pharmacovigilance, such as uncovering medication side effects, adverse drug reactions, and off-label usage trends from unstructured data to support large-scale drug safety monitoring efforts, and in healthcare education and training, where ENACT NLP can provide valuable case studies by extracting real-world clinical scenarios from narratives, enhancing training programs and AI tool development through exposure to authentic patient data. Collectively, these applications expand ENACT NLP’s impact, offering powerful solutions for improving healthcare research, policy, and practice.

### Lessons Learned

Implementing and deploying NLP infrastructures within the ENACT network has been a multifaceted journey, marked by significant advancements in integrating NLP and textual analytical capabilities into a large national-scale EHR network. The ENACT NLP Working Group’s collaborative efforts, leveraging the existing IT infrastructures and NLP expertise at various CTSA hubs, have facilitated the rapid deployment of NLP pipelines across the network. Establishing a Slack workspace and other communication channels was instrumental in ensuring smooth coordination and troubleshooting among participating sites. The experiences garnered through this process underscore the importance of a well-coordinated, collaborative approach in deploying complex, multisite informatics solutions.

Despite the successes, the implementation process encountered several notable challenges. A primary obstacle was the sheer volume of unstructured clinical notes available at each site, which made processing all notes and extracting every relevant medical concept impractical. Furthermore, developing a generalized NLP algorithm capable of handling all possible medical concepts across diverse clinical contexts proved unrealistic, leading us to take a more realistic approach in multiple focus groups. The limited funding allocated to the project also restricted the number of NLP algorithms that could be developed and comprehensively evaluated, necessitating a focus on specialized algorithms for specific projects. Additionally, the variability in data types and note formats across different sites introduced complexities in standardizing and harmonizing the NLP-derived data. Some participating sites have successfully tested the process using synthetic data and are now transitioning to working with a gold-standard dataset. However, the implementation timelines differ across sites due to variations in personal experience and availability and delays in IRB approvals.

During the implementation and deployment of the NLP within the ENACT network, we didn’t use any theoretical framework to guide the implementation process. However, implementation science^35,36^, a discipline commonly used in intervention research and public health programs, could potentially guide the practical implementation and deployment of NLP and other artificial intelligence (AI) models in complex, diverse real-world settings. Implementation science offers many valuable frameworks, such as the Exploration, Preparation, Implementation, Sustainment Framework (EPIS)^37,38^ and The Consolidated Framework for Implementation Research (CFIR)^37^. By systematically studying the factors that influence the successful adoption and integration of these models, implementation science can provide insights into optimizing workflows, enhancing collaboration among stakeholders, and overcoming barriers to scalability. For instance, implementation science could help develop tailored strategies to adapt NLP algorithms to the specific needs and constraints of different sites, thereby improving the overall effectiveness and sustainability of the deployment. Moreover, it can facilitate the identification of best practices for scaling AI-driven solutions across diverse healthcare environments, ultimately accelerating the translation of NLP-derived insights into clinical practice.

### Limitations

The process described in this paper has several limitations that warrant consideration. First, the focus on specific projects and the development of specialized NLP algorithms, while necessary due to resource constraints, may limit the generalizability of the findings to other clinical contexts. NLP models trained and validated on certain datasets may not perform as effectively on different patient populations, specialties, or healthcare settings, requiring additional adaptation and validation efforts. Additionally, the variability in data quality, note types, and EHR systems across the participating sites poses challenges in ensuring consistent performance of the NLP algorithms. Differences in documentation practices, clinical terminologies, and system configurations could introduce inconsistencies that affect model robustness and accuracy, necessitating site-specific tuning. The reliance on local funding and existing funded projects for developing certain NLP tools also introduces potential biases, as the algorithms may be optimized for specific datasets that are not representative of the broader population. This funding-driven approach may inadvertently prioritize projects with greater institutional support while leaving gaps in NLP capabilities for underrepresented patient groups and clinical domains.

Third, the querying function across the network is still under development, limiting the immediate utility of NLP-derived fields for large-scale, multi-site research. Some sites have onboarded onto the ENACT test network to assist in refining the querying capabilities, but due to the complexity of each site’s infrastructure and variations in personnel bandwidth, progress has been gradual. We anticipate that the querying function will become available to sites within the working group by mid-2025 and will be extended across the entire ENACT network by late 2026. The ability to effectively query NLP-derived data is crucial for integrating unstructured text into clinical and translational research, and continued investment in this functionality is necessary.

Finally, the lack of comprehensive evaluation across all ENACT sites limits the ability to fully assess the scalability and adaptability of the deployed NLP infrastructures. While individual sites have conducted rigorous validation, cross-site generalizability remains uncertain. Differences in patient populations, clinical documentation styles, and computing environments could introduce unforeseen challenges when deploying NLP models at scale. Future work should focus on expanding evaluation efforts across diverse sites, implementing federated validation frameworks, developing evaluation tools and visualization dashboards, and exploring strategies to mitigate site-specific biases. Additionally, ongoing collaboration with clinicians and informaticians will be essential to refine NLP methodologies, enhance usability, and ensure meaningful clinical impact.

### Generative AI and LLMs in ENACT NLP

Recent advancements in generative AI (genAI) and large language models (LLMs) can potentially address several key limitations in the ENACT NLP project. First, current limitations in developing generalized NLP algorithms across diverse health systems could be alleviated using LLMs. Unlike specialized NLP algorithms, LLMs such as GPT-4 can be fine-tuned to understand clinical context across various datasets without requiring domain-specific rules. This generalization ability could help ENACT develop more versatile NLP tools to handle multiple clinical tasks (e.g., phenotyping, cohort identification) across different sites without extensive retraining. Second, LLMs could enhance the accuracy of phenotyping efforts, particularly in multi-site studies, where heterogeneity in data sources makes consistent concept extraction difficult. Generative AI models, especially those trained on clinical data, can significantly improve this task by capturing nuanced clinical language. LLMs can interpret complex medical narratives more effectively than rule-based systems and adapt to new or evolving medical terminologies. As an example, open source LLMs (e.g., Llama2-70B-chat, Openchat-3.5-0106) were able to identify mammograms that required follow up with F1 = 1.0 (i.e., perfect performance in that experiment) based on text reports. Notably, mammography reports include a Breast Imaging-Reporting and Data System (BI-RADS) score. A mammogram (report) that requires follow up is one where the interpreting radiologist assigned a BI-RADS score other than 1 or 2. Thus, identifying mammograms that require follow up is a relatively simple information extraction task^39^. Third, LLMs may reduce the time to develop NLP algorithms. LLMs bring the advantage of being pretrained on diverse datasets, allowing them to incorporate time-consuming external knowledge into knowledge engineering in traditional rule-based NLP systems. Fourth, LLMs could automate multisite validation and deployment. One of the key bottlenecks for ENACT is the complex logistics of multisite validation of NLP tools. GenAI models could streamline this by providing automated validation.

While genAI and LLMs offer considerable potential to advance the ENACT NLP initiative, several significant challenges and drawbacks must be considered. First, data privacy and security concerns are paramount in healthcare, as LLMs typically require large amounts of data to train and fine-tune. This presents the risk of inadvertently exposing sensitive patient information, especially when models are trained across multiple sites in a distributed network like ENACT. Even anonymized or de-identified data may still contain subtle information that could re-identify individuals, posing a significant risk under regulations such as HIPAA. Additionally, the computational and resource costs associated with training, fine-tuning, and deploying LLMs are substantial, and for a large, multisite initiative like ENACT, these infrastructure costs could be prohibitive for sites with limited resources, leading to disparities in model performance and inconsistent results across the network. Another concern is bias, fairness, and ethical considerations, as LLMs often inherit biases from their training data. In healthcare, biased models could disproportionately affect certain demographic groups, leading to incorrect or harmful clinical recommendations. This is particularly problematic in NLP tasks such as phenotyping or clinical decision support, where subtle biases in language or data representation could skew interpretations. The “black box” nature of LLMs also poses challenges in clinical applications where interpretability is crucial, as clinicians and researchers often need to understand why a model made a particular prediction. This lack of explainability can lead to trust issues in the healthcare domain. The ethical implications of using genAI in healthcare, particularly LLMs, are still evolving. Using LLMs raises numerous ethical and legal questions, from patient consent to data ownership and the responsibility for errors or adverse outcomes. These issues are particularly complex in a multisite network like ENACT, where multiple stakeholders may be involved in data sharing, algorithm development, and model deployment. Moreover, LLMs sometimes generate plausible-sounding but incorrect information, a phenomenon known as “hallucination.” This could have serious consequences in clinical contexts, leading to misinformed decisions based on incorrect data extraction, summarization, or interpretation of clinical narratives. While GenAI and LLMs hold promise for advancing the work of ENACT NLP, the challenges surrounding their implementation, especially in areas like privacy, bias, explainability, and ethics, must be carefully managed. A balanced approach combining the power of LLMs with traditional rule-based methods and rigorous oversight may offer the best path forward for ENACT NLP objectives.

## Conclusion

The ENACT NLP Working Group has made significant strides in deploying NLP infrastructure across a large, federated data network, leveraging both existing IT infrastructure and NLP expertise from several CTSA hubs. By establishing focus groups dedicated to specific disease conditions and utilizing the Open Health NLP (OHNLP) Toolkit, the Working Group was able to target specialized NLP algorithms for distinct clinical tasks. This pragmatic approach has enabled the ENACT network to deploy NLP solutions more efficiently while addressing each site’s unique data and resource challenges. Furthermore, the collaborative framework of partnerships within the OHNLP development team has been crucial in facilitating rapid implementation and troubleshooting.

The project has faced challenges in processing vast amounts of clinical notes and in developing NLP algorithms that can perform consistently across all sites. The Working Group opted for focused NLP deployments and clearly defined cohort specifications to address these obstacles, tailoring algorithms to specific note types and clinical contexts. As the project evolves, creating and refining these NLP tools, as well as emphasizing collaboration and resource-sharing, will be critical in broadening the scope and impact of the ENACT NLP initiative across diverse healthcare environments.

## Methods

### NLP Tools

We chose the Open Health Natural Language Processing (OHNLP) Toolkit^19^, developed by the OHNLP consortium, as our primary NLP tool for extracting entities from clinical notes. The OHNLP Toolkit employs an ontology-driven, dictionary-based approach with MedTagger^20^ as the core entity extraction component. The Toolkit is highly adaptable, accepting input from local file systems, relational databases, and the Epic Clarity reporting database and delivering output to file systems and relational databases. Compared to other NLP tools like MetaMap^21^, the Toolkit offers higher throughput, better customization, and increased scalability, making it ideal for processing large volumes of notes. Its effectiveness has been demonstrated in large EHR data projects like the National COVID Cohort Collaborative (N3C)^22^. Furthermore, we partnered with the OHNLP development team and created a dedicated support channel on Slack to assist sites with troubleshooting during validation and deployment.

The OHNLP Toolkit is built using the Java programming language. Recognizing that some sites do not use Java, we are developing a Python alternative that uses SpaCy^23^ and can be deployed on servers running the Linux, macOS, and Windows operating systems. Like the Toolkit, the Python alternative will accept input from various sources, including file systems, .csv files, .zip archives, and relational databases, and deliver output to file systems and relational databases. Another key feature is that it will be compatible with the Toolkit rulesets, which enables smooth switching between the two tools without losing functionality. We plan to make the Python alternative publicly available through the OHNLP consortium’s GitHub repository^24^, ensuring it is freely available to the informatics and biomedical research communities.

### Extension of the Common Data Model for NLP-derived Data

The ENACT network uses the ACT/i2b2 common data model (CDM)^25^ to harmonize the EHR data (more recently, the OMOP CDM has been compatible with ENACT; in the future, the PCORnet CDM will also be compatible). In the ACT/i2b2 CDM, a concept is a standardized term from a medical terminology (e.g., ICD-10-CM) or a custom terminology (e.g., a hospital’s in-house oncology terminology), and a fact is a particular instance or observation of a concept for a specific patient (e.g., ICD-10-CM:S92.4). NLP-derived entities or facts are stored in the i2b2 *observation_fact* table, which also stores structured EHR data. Detailed documentation on concepts and facts and their representations in the *observation_fact* table are available at the i2b2 Wiki^26^. The Working Group established a set of conventions for representing NLP-derived entities in the *observation_fact* table outlined in Table 2. The conventions are grounded in a philosophy emphasizing scalability and customization to meet the diverse needs of sites and projects. Recognizing that each site may have unique requirements and each project may include differing granularity of NLP-derived data, the conventions are designed to be flexible, allowing for adjustments and extensions as needed. By prioritizing adaptability, these conventions support the broad applicability and long-term sustainability of NLP in the ENACT network.

The CONCEPT_CD field in the *observation_fact* table is the key field to store NLP-derived entities. Because structured EHR data is also stored in the *observation_fact* table, it is critical to distinguish NLP-derived data from them. To accomplish this, NLP-derived entities are prefixed with the string “NLP.” NLP-derived entities are translated to concepts obtained from standardized medical terminologies such as the ICD-10-CM or a project’s custom terminology. When using a concept from a standard terminology, the entry in the CONCEPT_CD field in the *observation_fact* table is formatted as NLP|<PROJECT_NUM>|<STANDARD_VOCABULARY_PREFIX>:<STANDARD_CONCEPT_CODE>, where <STANDARD_VOCABULARY_PREFIX> indicates the standard terminology used and <STANDARD_CONCEPT_CODE> represents the concept in the standard terminology. For example, the entity “fracture of a great toe” extracted by an NLP algorithm is represented as NLP|001|ICD-10-CM:S92.4 in the CONCEPT_CD field. When using a concept from a custom terminology, the entry in the CONCEPT_CD field is formatted as NLP|<PROJECT_NUM>|CUSTOM|<PROJECT_NAME>:<CONCEPT>, where <PROJECT_NAME> specifies the project, and <CONCEPT> describes the custom concept. For example, the entity “snoring” extracted by an NLP algorithm in a project on sleep is represented as NLP|001|CUSTOM|SLEEP:SNORING in the CONCEPT_CD field. **Table 5** provides additional examples.

**Table 5.**
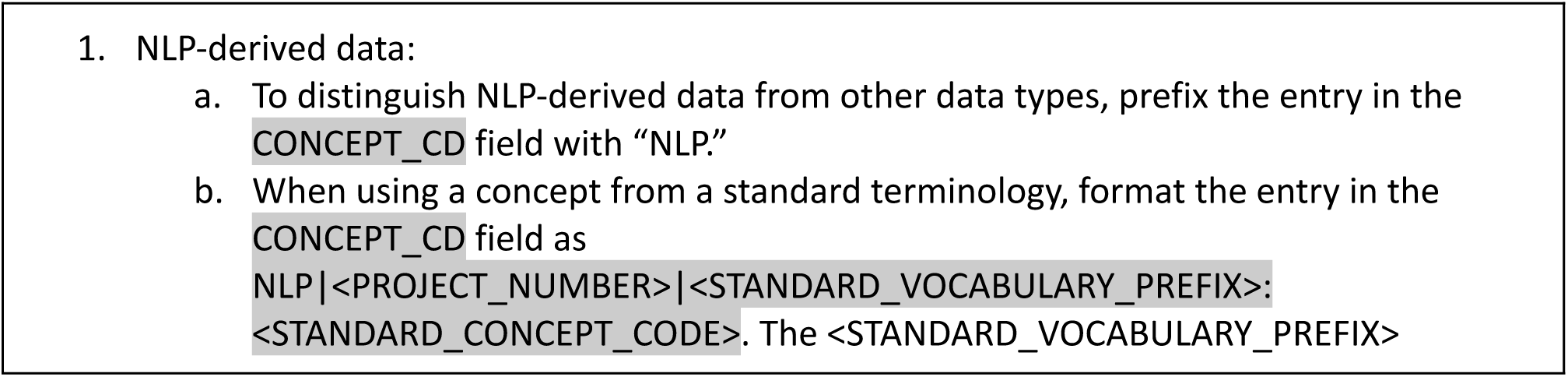
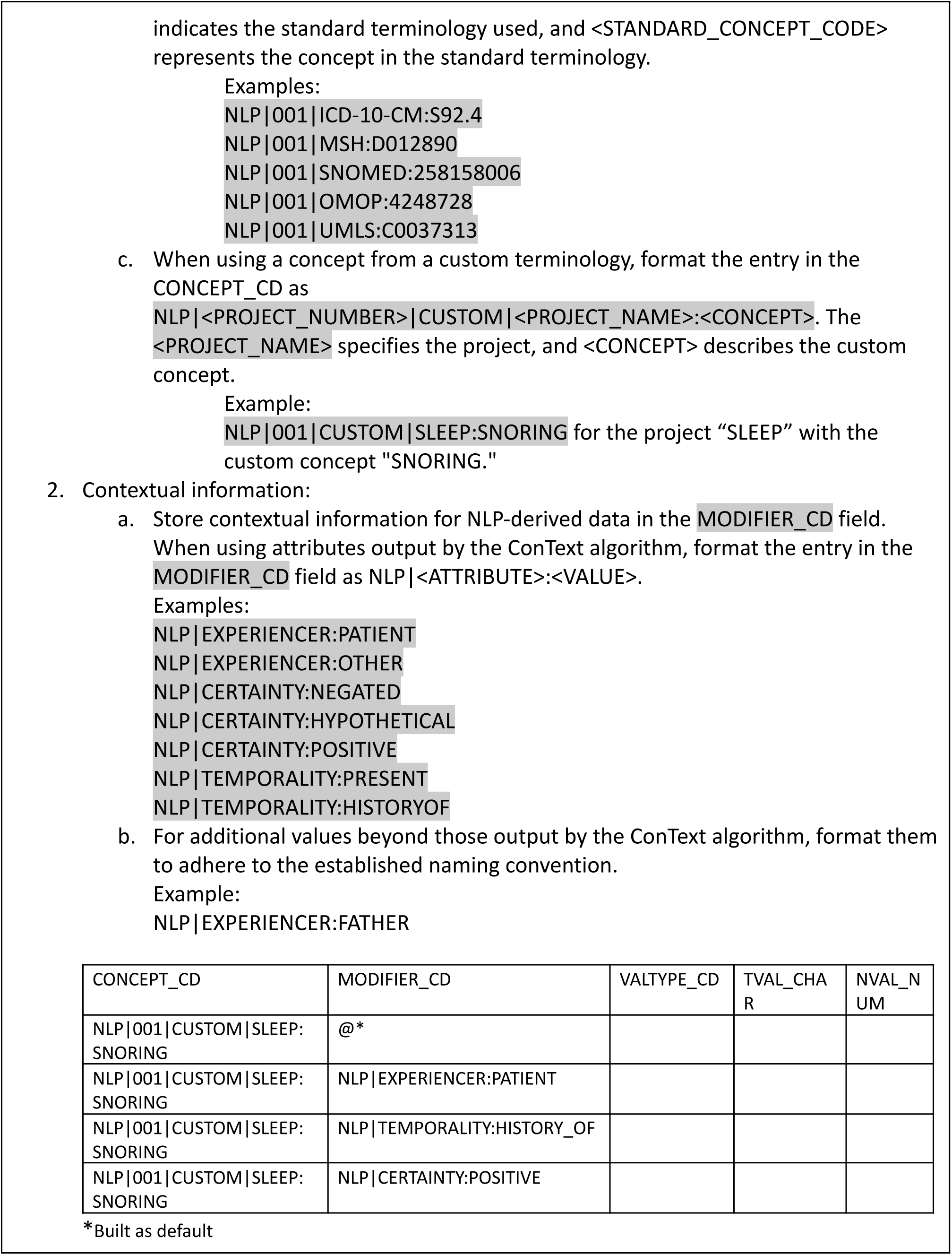
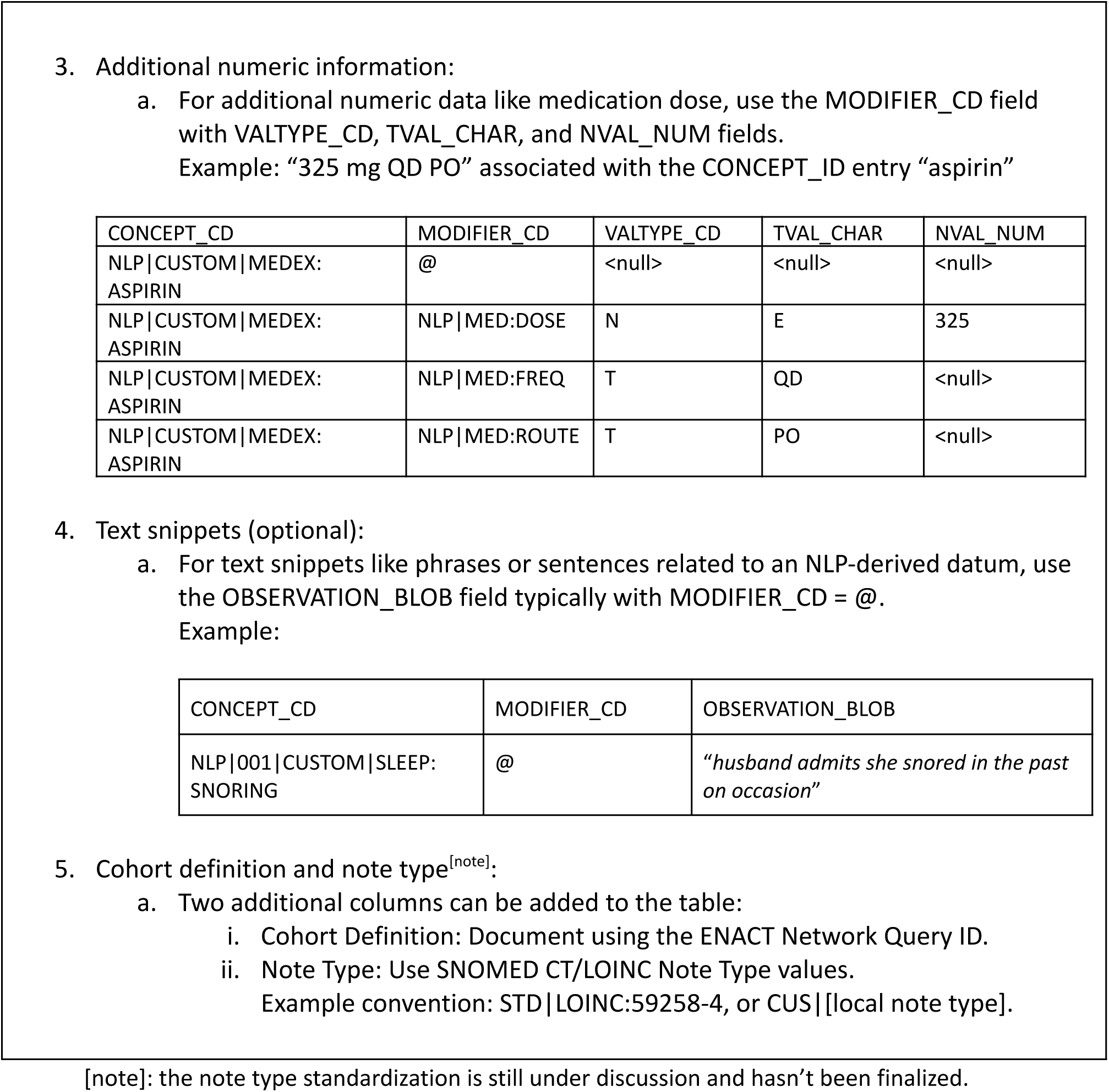
Conventions for using i2b2 *observation_fact* table for NLP-derived data.

In addition to the main entity, NLP algorithms often output contextual information, with most using the ConText algorithm^27^, which includes attributes such as *experiencer*, *certainty*, and *temporality*. The OHNLP Toolkit also employs the ConText algorithm to extract contextual information. The MODIFIER_CD field in the *observation_fact* table is used to store contextual attributes, formatted as NLP|<ATTRIBUTE>:<VALUE> (e.g., NLP|EXPERIENCER:PATIENT). A single NLP-derived entity may be associated with multiple contextual attributes, requiring multiple MODIFIER_CD entries. This could cause the *observation_fact* table to become very large, potentially consuming significant storage. However, this limitation is offset by the enormous flexibility of the entity-attribute-value table design of i2b2, allowing it to store any type of NLP-derived data without needing custom database tables. For example, an NLP-derived medication entity may include contextual attributes related to dose. The MODIFIER_CD field, together with VALTYPE_CD, TVAL_CHAR, and NVAL_NUM, is used to store numeric attributes such as “325 mg QD PO” associated with the CONCEPT_ID entry “aspirin” (see Table 2).

Sometimes, text snippets like phrases or sentences related to an NLP-derived entity may be useful for research; such text snippets are stored in the OBSERVATION_BLOB field in the *observation_fact* table. However, populating text snippets is optional since they may inadvertently contain protected health information, and ENACT current policy allows only limited datasets in the network’s data repositories. Beyond phrases or sentences, other types of information, such as location data, may also be stored in the OBSERVATION_BLOB field.

The Working Group has proposed the addition of two new columns to the *observation_fact* table to store the cohort definition and note type. A key benefit of the ENACT network is the consistent definition of cohorts since each cohort definition query is associated with a unique Query ID propagated throughout the network. This unique Query ID can unambiguously identify the cohort on which an NLP algorithm was run. Standard terminologies for note type are available, for example, in SNOMED CT and LOINC (e.g., STD|LOINC:59258-4 denotes a standard document type according to LOINC Note Type). Local custom terminologies may also be used (e.g., CUS|[local note type]). For a project, sites typically agree on the specific note type from which to extract NLP data.

### Extension of the Ontology for NLP-derived Data

To query the NLP-derived data in the ENACT network, the Working Group has extended the ACT ontologies to include concepts that represent NLP-derived entities. The ontologies for NLP-derived data are visually separated from the existing ones for structured EHR data in the SHRINE interface. Two new ontologies have been developed. The first ontology, Note Types, allows identifying types of clinical notes and is based on LOINC’s document ontology. The current LOINC document ontology describes key attributes of clinical documents in five axes: type of service, kind of document, setting, role, and subject matter domain, with each axis having a set of controlled terms. The second ontology, Clinical Concepts by Project, provides project-specific concepts for querying task-specific NLP-derived entities. Each project-specific ontology is provided as a separate subtree, and concepts can be derived from standard terminologies such as ICD-10 or a custom terminology developed for the project. For example, the Rare Disease ontology contains concepts from the Orphanet Rare Disease Ontology (ORDO), while the Sleep ontology uses custom concepts like DAY_SLEEP and SLEEP_PROBLEMS. Figure 3 provides a screenshot of the current ENACT NLP ontologies.

**Figure 3.**
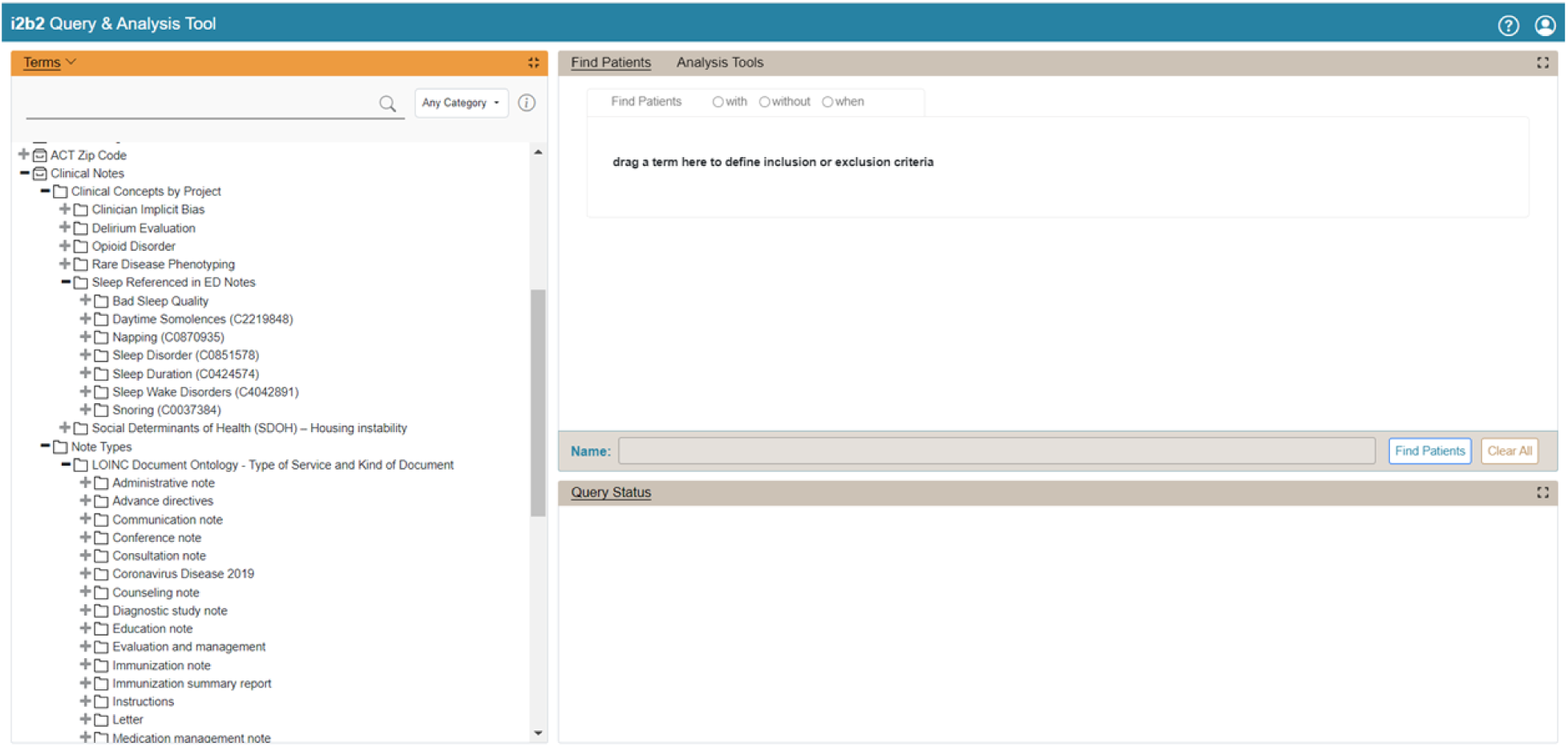
ENACT NLP ontology implementation.

Whenever possible, project-specific ontologies are designed to leverage standard vocabularies to enable reuse and compare entities derived from both structured EHR data and clinical notes. Contextual information stored in the CONCEPT_CD field and additional information stored in the OBSERVATION_BLOB field are currently not queryable through the SHRINE interface, but data stored in the CONCEPT_CD field can be queried via the i2b2 interface.

### Evaluation of NLP Algorithms

We developed an approach for evaluating NLP algorithms that accommodates the federated nature of the ENACT network, where clinical notes from multiple institutions cannot be aggregated at a central location. Consequently, the evaluation framework emphasizes intra-site and cross-site validation, facilitated by explicitly shared definitions of cohort specification and applicable note type, a gold standard that development sites manually derive, and a computable silver standard that requires minimal manual review by deployment sites. Figure 4 provides an overview of this approach

**Figure 4.**
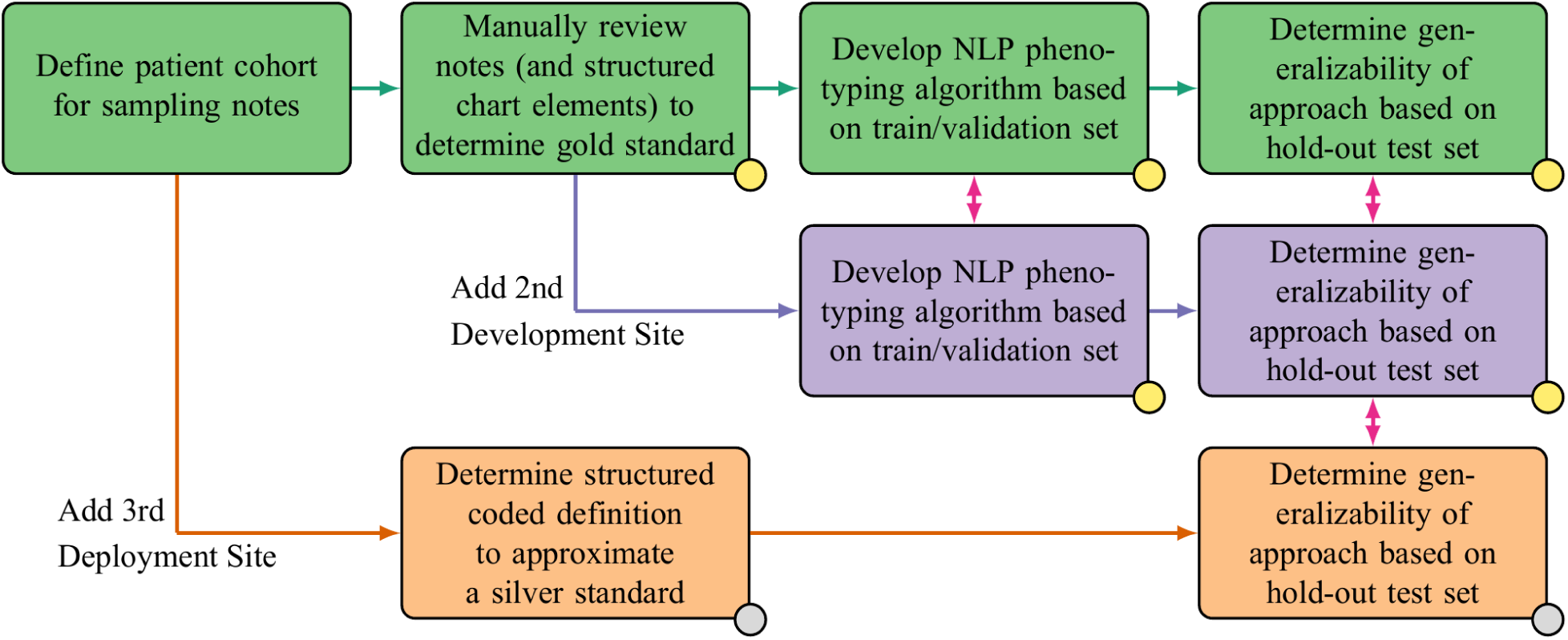
Evaluation of NLP alogrithms across development and deployment sites in the national ENACT network. The green, purple, and orange boxes indicates tasks undertaken at each of the three types of sites, respectively. The tasks associated with a gold standard have a gold circle and the tasks associated with a silver standard have a silver circle. The red arrows between sites indicate the possibility of a cross-site evaluation or comparsion.

Specifications for an algorithm start with a development site that defines a cohort, an applicable note type, and a method for sampling the notes when a large volume is available. A second development site reviews and adapts these specifications for generalizability; for example, they may clarify the definition of the note type and the sampling procedure. Once the specifications are finalized, the development sites employ a manual review process to establish a gold standard reference for the NLP algorithm. In conjunction with this gold standard reference, the sites mutually agree upon a silver standard reference, which can be derived from structured data alone or with minimal additional manual review. The development sites iteratively refine the algorithm, allowing for comparisons of rules, models, and, most critically, unforeseen challenges.

Once the algorithm development is completed, it is shared with the Working Group and the validation sites. In rare cases, the algorithm cannot be shared among sites due to privacy concerns; in such cases, the detailed training method is provided, allowing each site to construct the algorithm using its own data. The development sites assess the algorithm using both gold and silver standard references. They compare the performance of these two standards to ensure that the silver standard is adequate for broader evaluation at other sites. Finally, the validation sites test the algorithm at each of their locations using the readily computable silver standard reference.

This evaluation approach lowers the barrier to cross-site validation of NLP algorithms by establishing a consistent and replicable framework for effective collaboration among sites. The workflow shown in Figure 3 provides a clear path for sites to evaluate algorithms in their own environments, which may differ somewhat from the environments at the development sites. This approach minimizes the need for extensive manual interventions, accelerates the validation process, and promotes broader adoption of NLP algorithms across multiple sites. As cross-site validation becomes increasingly efficient, streamlined, and scalable, NLP algorithms can be feasibly deployed across the entire ENACT network.

### Standardization of Error Analysis

Error analysis is a common practice in developing and validating NLP algorithms. Figure 5 presents an overview of the site-level error analysis process adopted by the Working Group. We used our previously created error taxonomy and the clinical Text Retrieval and Use towards Scientific rigor and Transparent (TRUST) process^28^ to guide and standardize the error analysis. We focused on identifying the four most common error types: logic errors, linguistic errors, contextual errors, and annotation errors. We also gathered information about the task, the focus area (e.g., disease, medication class, or SDoH category), site expertise in clinical NLP, the NLP methodology, and annotation details. We utilized error analyses during algorithm development and applied the results to enhance performance, as well as after the algorithm’s development to collect data on its generalizability, portability, and limitations.

**Figure 5.**
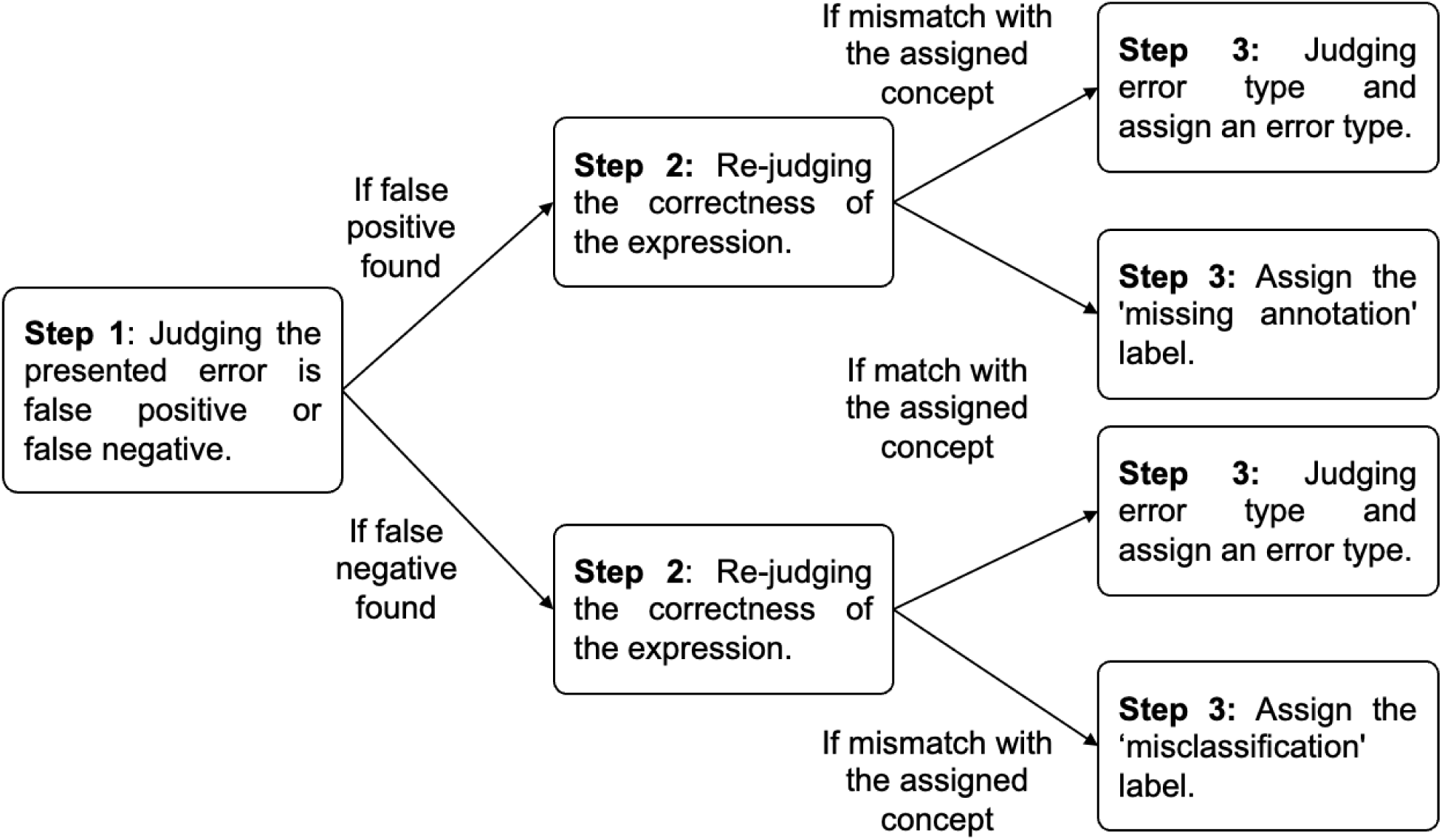
Overview of the logic and process for multisite error analysis.

## Data Availability

The patient-level electronic health record data used in this paper cannot be shared due to privacy and legal reasons.

## Code Availability

The codes utilized in this study are primarily open-source. The OHNLP Toolkit is available at OHNLP website (https://ohnlp.org/). The ENACT ontologies can be accessed at ENACT Network Resources (https://www.enact-network.us/resources/technical). Documentation for the i2b2 Common Data Model (CDM) is available at i2b2 Wiki (https://community.i2b2.org/wiki/display/BUN/i2b2+Common+Data+Model+Documentation).

## Acknowledgements

The research reported in this publication was supported by the National Institutes of Health under award numbers U01TR002062, UL1TR001998, UM1TR004906, U01TR002628, UL1TR003163, UL1TR001857, U24TR004111, 30TR002103, UL1TR002001, UL1TR002377, and UM1TR004407 from the National Center for Advancing Translational Sciences, award numbers RF1AG072799, R01AG060993, R01AG077017, and R01AG068007 from the National Institute on Aging, award number R01LM014306 from the National Library of Medicine, award number R01GM141476 from the National Institute of General Medical Sciences, and the Reynolds and Reynolds Foundation. The content is solely the responsibility of the authors and does not necessarily represent the official views of the funding agencies.

## Author Contributions

J.H. and Y.W. conceptualized, designed, and organized this study, analyzed the results, and wrote, reviewed, and revised the paper. Y.W., J.H., C.L., M.M., P.M.H., S.F., M.J.K., A.W., J.R.A., L.W., E.B., H.L., J.C., D.R.H., A.C., D.H., J.D.O., R.E.K., N.E.G.R., J.F.R., C.Y., Y.C., M.B.P., T.J.M., B.A.M., C.M.P., J.W.F., S.S., S.P., Y.P., A.P., Y.W., Z.X., S.L., S.E.R., and S.V. wrote, reviewed, and revised the paper. Y.W. and S.V. conceptualized, designed, and directed this study, wrote, reviewed, and revised the paper.

